# Anosmia and dysgeusia in patients with mild SARS-CoV-2 infection

**DOI:** 10.1101/2020.04.11.20055483

**Authors:** Ruth Levinson, Meital Elbaz, Ronen Ben-Ami, David Shasha, Tal levinson, Guy Choshen, Ksenia Petrov, Avi Gadoth, Yael Paran

**Affiliations:** Internal medicine clinic, Tel Aviv Sourasky Medical Center, Tel Aviv, Israel; Infectious Diseases Unit, Tel Aviv Sourasky Medical Center, Tel Aviv, Israel; Internal Medicine H, Tel Aviv Sourasky Medical Center, Tel Aviv, Israel; Clinical Research Unit, Oncology Division, Tel Aviv Sourasky Medical Center, Tel Aviv, Israel; Department of neurology, Tel Aviv Sourasky Medical Center, Tel Aviv, Israel; Sackler Faculty of Medicine, Tel Aviv University, Tel Aviv, Israel

**Keywords:** SARS-CoV-2, COVID-19, Anosmia, Dysgusia

## Abstract

In this cohort of 42 patients with mild COVID19 we found a unique clinical feature of acute anosmia and dysgeusia in more than third of patients. Median onset of these features was 3.3 days after onset of illness (range 0-7) with rapid recovery in most patients.

**Article Summary Line:** Anosmia and dysgeusia appeared early in third of our patients with mild SARS-CoV-2 infection and were short-lived in most patients.

## Introduction

On March 10, 2020 an isolation unit for patients with mild coronavirus disease (COVID)-19 was opened at the Tel Aviv Medical Center (TASMC), Israel. Within the first week, we noted that many patients reported complete loss of sense of smell (anosmia) as well as altered taste sensation (dysgeusia), unrelated to appetite or nasal congestion symptoms. Anecdotally, patients described being unable to smell a lemon under their nose, or that every food they ate tasted like paper or saliva.

Here, we aimed to describe the prevalence, time of onset in relation to disease onset, and duration of anosmia and dysgeusia.

## Methods

We reviewed the symptoms of hospitalized adults and adolescents (age ≥15 years) admitted to the isolation unit with COVID-19 between March 10-23, 2020. We included patients with positive PCR for SARS-CoV-2 from naso-pharyngeal swabs and mild symptoms, defined as National Early Warning Score (NEWS) 0 to 5 (*1*). All patients were symptomatic.

Demographic data, comorbid conditions, clinical symptoms and signs were extracted from the hospital electronic medical record system.

In addition, patients were asked to complete a questionnaire administered via mobile phone or email. A phone interview was conducted after discharge every 3-5 days until resolution of anosmia and dysgeusia, or up until 25^th^ of March. The study was reviewed and approved by the TASMC ethics committee (approval number TLV-0206-20). Patients were asked to give their consent to participate in the study by phone. A waiver from signed informed consent was given by the ethic committee.

The first day of illness was defined as the day of onset of the first related symptom.

## Results

### Study population

Forty-five patients were admitted to the isolation unit between March 10-23, 2020, all with mild COVID-19 (NEWS score 0 to 5 at peak illness). All patients were hospitalized for observation and isolation, and none required medical therapy or supplemental oxygen. Forty-two patients (93%) completed the questionnaire and were included in our analysis. Median age was 34 years (range 15-82 years), 23 males and 19 females. 6 (9.5%) had comorbid medical conditions and 4 (4.8%) had a history of smoking (Table S1).

Anosmia, dysgeusia and other symptoms

Fifteen patients (35.7%) reported anosmia and 14 (33.3%) reported dysgeusia. Fourteen patients reported both anosmia and dysgeusia, and one reported only anosmia.

Anosmia and dysgeusia started a median of 3.3 days after illness onset (range, 0 to 7 days). By the time of writing, 11 of 15 patients (73.3%) had reported recovering both their sense of smell and taste. The median duration of these symptoms was 7.1 days for dysgeusia (range, 0-7 days) and 7.6 days for anosmia (range, 4-17 days) (Figure 1). Of the remaining 4 patients, 2 reported recovering only sense of taste with ongoing anosmia for 14 and 17 days, respectively, and 2 reported ongoing anosmia and dysgeusia for 11 and 14 days, respectively.

**Figure 1:**
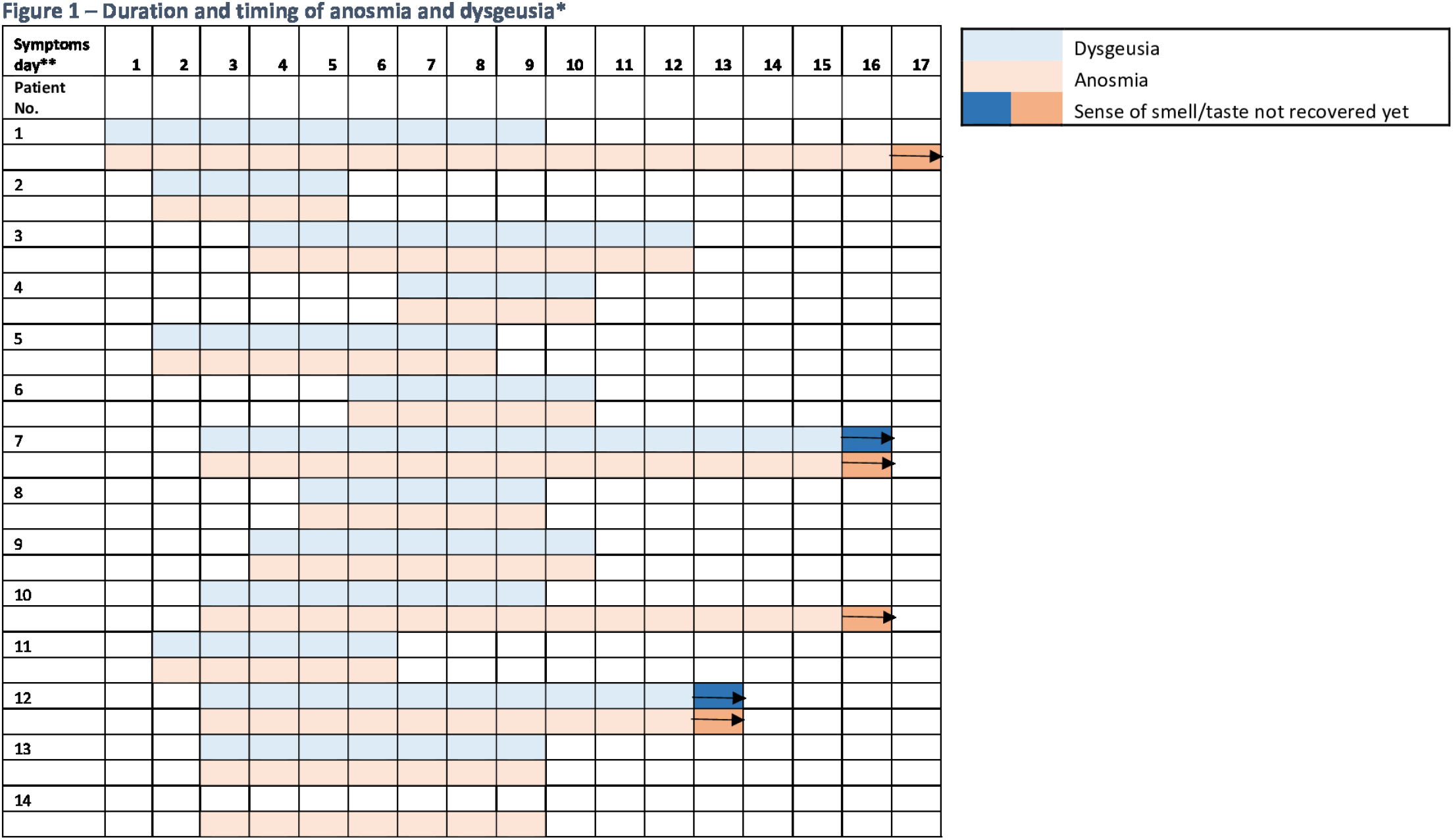
Duration and timing of anosmia and dysgeusia in 14 patients. * One patient whose symptoms lasted for 5 days is not shown since start of symptoms is unknown. ** Day 1 is the day of the first symptom during illness.

Anosmia and dysgeusia were not associated with age, sex, BMI or any comorbid condition, including smoking. However, the duration of anosmia, but not dysgeusia, was significantly longer for patients older than 40 years of age (mean duration, 11.5 days versus 6.1 days for patients younger than 40 years; P=0.01).

Prevalence of other symptoms during the course of illness is shown in supplementary Table2. The most common symptoms were cough (69%), fatigue (69%), fever ≥ 38.0° C (66.6%) and myalgia and/or arthralgia (57%). Nine patients (21.4%) had diarrhea and 7 (16.67%) had abdominal pain.

Anosmia and dysgeusia were more frequent in patients with sore throat (anosmia, 69.2% versus 20.7%; P=0.005 ; dysgeusia, 61.5% and 20.7%; P=0.015) and anosmia was more frequent in patients with dyspnea (55.6% and 20.8%; P=0.02). Anosmia and dysgeusia were not associated with other symptoms, including coryza.

## Discussion

Anosmia and hyposmia, the inability or decreased ability to smell, is estimated to afflict 3%–20% of the population, increasing in frequency with age (*2*). Patients often have difficulty discriminating between smell and taste dysfunction (*3*). Moreover, these common disorders are often overlooked by medical professionals (*3, 4*). Acute onset anosmia usually follows viral infections or trauma (*5*). Upper respiratory tract infection is one of the most commonly identified causes of olfactory loss, accounting for 22% to 36% of cases(*6*).

We found that anosmia or dysgeusia are common in patients with mild COVID-19 and were self-reported by more than a third of patients. In contrast to post-viral anosmia and dysgeusia reported with common cold or influenza, symptoms appeared early (median of 3.3 days) after COVID-19 onset, rather than after illness resolution (*7*). Moreover, anosmia and dysgeusia were short lived and resolved within approximately 7 days in most patients with COVID-19, whereas recovery time is typically several weeks to months for other post-viral diseases (*8, 9*). Patients younger than 40 years recovered olfaction more rapidly than patients older than 40 years.

The mechanism of smell disorder in post-viral anosmia is believed to be viral damage to the olfactory epithelium independent of nasal congestion (*2*). The volume of the olfactory bulbs is known to be decreased in post-infectious olfactory dysfunction and correlates with olfactory function and even serve as prognostic factor in these patients (*10*). In a recent case report by Eliezer et al (*11*) a patient with COVID-19 and complete olfactory loss had a bilateral obstructive inflammation of the olfactory clefts on imaging without other nasal obstruction or rhinorrhea, and no anomalies of the olfactory bulbs and tracts. This upper obstruction presumably prevented odorant molecules from reaching the olfactory epithelium.

These findings are compatible with our results, showing no association between anosmia or dysgeusia and coryza. In fact, fewer than half the patients with these symptoms had coryza.

The incidence of postinfectious smell dysfunction associated with coronaviruses other than SARS-Cov-2 has been reported only in a limited number of studies (*12*).

COVID-19 emerged in Wuhan, China, in November 2019 and has rapidly became pandemic. Early onset anosmia/dysgeusia are unique clinical features that might help differentiate COVID-19 from other common respiratory viral infections, such as influenza. This is of particular importance in primary care and in resource-limited settings, where rapid molecular diagnostic testing may not be readily available.

In the realty of a pandemic which infected more the 1.5 million people worldwide and the limitation in virologic confirmation test, familiarity with a unique symptom like this could help the primary physician in triaging which patients should be referred to virologic testing by naso pharyngeal swabs.

## Data Availability

all data are in the manuscript.

## Acknowledgments

None

## Biographical Sketch

Dr Ruth Levinson is a specialist in internal medicine and a senior physician at the Internal Medicine Consultation Department of the Tel Aviv Sourasky Medical Center. She is currently heading the Special COVID-19 Isolation Unit at the same institution. Her interests include diagnosis of complex medical cases and, more recently, researching the clinical presentation and natural history of COVID-19.

---

**Table 1.** Prevalence of Symptoms:

| Symptom | No. of Patients<br>with the<br>symptom | Prevalence (%)<br>Total N=42 |
| --- | --- | --- |
| Cough | 29 | 69 |
| Fatigue | 29 | 69 |
| Fever per History 37.9-37.9 | 9 | 21 |
| Fever per History $\geq 38.0$ | 28 | 67 |
| Myalgia or arthralgia | 24 | 57 |
| Headache | 20 | 48 |
| Coryza | 17 | 40 |
| Dyspnea | 18 | 43 |
| Anosmia | 14 | 33 |
| Dysgeusia | 15 | 36 |
| Sore Throat | 13 | 31 |
| Dizziness | 9 | 21 |
| Diarrhea | 9 | 21 |
| Abdominal pain | 7 | 17 |
| Nausea or vomiting | 5 | 12 |
| Palpitation | 5 | 12 |
| Chills | 2 | 5 |
| Chest pain | 2 | 5 |
| hemoptysis | 1 | 2 |
| Conjunctivitis | 1 | 2 |

## Appendix material

**Supplementary Table 1:**
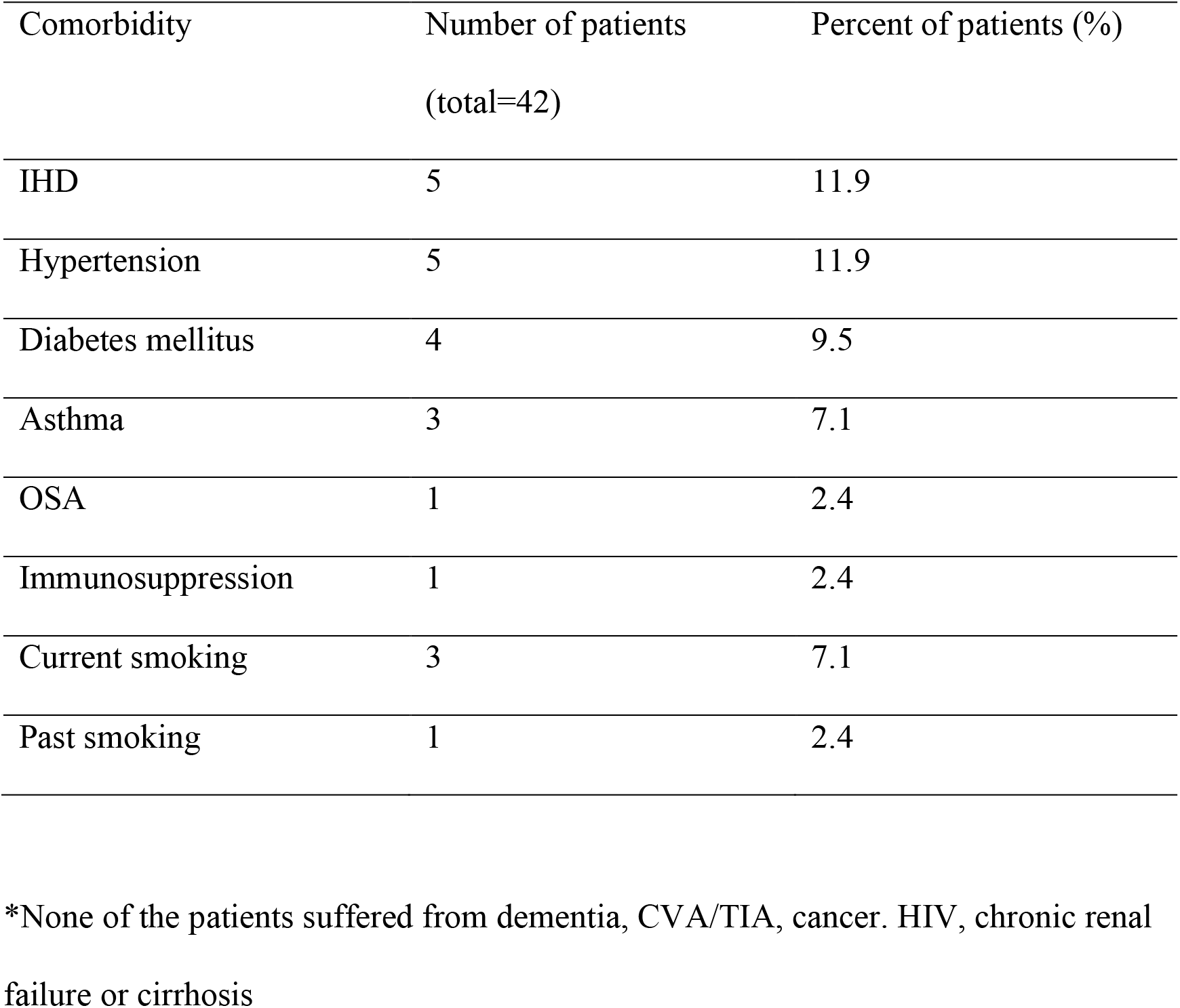
Comorbid medical conditions in patients with mild COVID-19

## References

1 Liao X, Wang B, Kang Y. Novel coronavirus infection during the 2019-2020 epidemic: preparing intensive care units-the experience in Sichuan Province, China. Intensive Care Med. 2020 Feb;46(2):357–60.

2 Hummel T, Rothbauer C, Barz S, Grosser K, Pauli E, Kobal G. Olfactory function in acute rhinitis. Annals of the New York Academy of Sciences. 1998 Nov 30;855:616–24.

3 Bromley SM. Smell and taste disorders: a primary care approach. American family physician. 2000 Jan 15;61(2):427-36, 38.

4 Schiffman SS. Taste and smell in disease (second of two parts). The New England journal of medicine. 1983 Jun 2;308(22):1337–43.

5 Boesveldt S, Postma EM, Boak D, Welge-Luessen A, Schopf V, Mainland JD, et al. Anosmia-A Clinical Review. Chemical senses. 2017 Sep 1.513–23:)7(42;

6 Seiden AM, Duncan HJ. The diagnosis of a conductive olfactory loss. The Laryngoscope. 2001 Jan;111(1):9–14.

7 Welge-Lussen A, Wolfensberger M. Olfactory disorders following upper respiratory tract infections. Advances in oto-rhino-laryngology. 2006;63:125–32.

8 Lee DY, Lee WH, Wee JH, Kim JW. Prognosis of postviral olfactory loss: follow-up study for longer than one year. American journal of rhinology & allergy. 2014 Sep-Oct;28(5):419–22.

9 Mori J, Aiba T, Sugiura M, Matsumoto K, Tomiyama K, Okuda F, et al. Clinical study of olfactory disturbance. Acta oto-laryngologica Supplementum. 1998;538:197–201.

10 Fonteyn S, Huart C, Deggouj N, Collet S, Eloy P, Rombaux P. Non-sinonasal-related olfactory dysfunction: A cohort of 496 patients. European annals of otorhinolaryngology, head and neck diseases. 2014 Apr;131(2):87–91.

11 Eliezer M, Hautefort C, Hamel AL, Verillaud B, Herman P, Houdart E, et al. Sudden and Complete Olfactory Loss Function as a Possible Symptom of COVID-19. JAMA otolaryngology--head & neck surgery. 2020 Apr 8.

12 Konstantinidis I, Haehner A, Frasnelli J, Reden J, Quante G, Damm M, et al. Post-infectious olfactory dysfunction exhibits a seasonal pattern. Rhinology. 2006 Jun;44(2):135–9.

